# Application of an EEG-based deep learning model to discriminate children with epileptic spasms from normal controls

**DOI:** 10.1101/2023.06.30.23292096

**Authors:** Mingjian Lu, Yipeng Zhang, Atsuro Diada, Shingo Oana, Rajsekar R. Rajaraman, Hiroki Nariai, Vwani Roychowdhury, Shaun A. Hussain

## Abstract

**Objective:** Given that epileptic spasms are often subtle, and that identification of hypsarrhythmia is limited by inadequate inter-rater reliability, there is a significant need for novel tools to aid the clinical identification of Infantile Epileptic Spasms Syndrome (IESS). Deep learning is an emerging technology which may enable efficient classification of disease states and may facilitate discovery of novel biomarkers. In this study, we set out to evaluate whether children with epileptic spasms can be distinguished from normal controls with use of an EEG-based deep learning model.

**Methods:** A deep learning model was trained and validated (5-fold cross validation) using 400 EEG samples (2 awake and 2 sleep samples from 50 children with epileptic spasms and 50 normal controls). Salient frequency bands and specific morphologic EEG features were identified with occlusion sensitivity analysis and targeted input perturbation, respectively.

**Results:** The model accurately distinguishes children with epileptic spasms from normal controls, solely on the basis of relatively short EEG samples. Using sleep data, accuracy = 0.95, recall = 0.96, precision (sensitivity) = 0.94, specificity = 0.94, and F1 score = 0.95. With awake data, accuracy = 0.91, recall = 0.84, precision = 0.98, specificity = 0.98, and F1 score = 0.90. The salient frequency bands for classification are 9.7 – 22.0 Hz and 1.0 – 6.8 Hz in sleep and awake EEG, respectively. With visual analysis of extracted salient features, we suspect that the model is identifying cases on the basis of paroxysmal fast activity in sleep and spike-wave activity in wakefulness.

**Conclusion:** This deep learning model represents a first step in the development of efficient algorithms that may aid in identification of epileptic spasms and IESS. More importantly, this approach may facilitate novel EEG-based biomarkers of epileptic spasms.

## Introduction

Infantile epileptic spasms syndrome (IESS) is a severe form of epileptic encephalopathy and the most common epilepsy syndrome in the first year of life.^1^ IESS begins in the first two years of life, and most often manifests with (1) epileptic spasms (ES; brief seizures often occurring in clusters), (2) hypsarrhythmia (a chaotic EEG pattern characterized by disorganized high-amplitude slowing and abundant epileptiform discharges), and (3) neurodevelopmental stagnation or regression.^2,3^ Although a majority of patients with IESS fulfill these criteria, there are many cases in which ES are exceptionally subtle, hypsarrhythmia is “mild”, or developmental impairment is initially absent or unrecognized. In addition, there are many cases in which ES occurs in older children, typically without hypsarrhythmia, and intermixed with other forms of epilepsy.^4^ The differential diagnosis chiefly consists of gastroesophageal reflux, benign sleep myoclonus, a spectrum of normal infant behaviors, and other forms of epilepsy. Given this milieu, recognition of ES and IESS is clinically challenging and requires extended video-EEG to electrographically evaluate both ictal events (which often only occur upon first morning awakening) and interictal hypsarrhythmia (which may be intermittent). However, even this gold-standard evaluation is problematic, in that the identification and characterization of hypsarrhythmia exhibits imperfect inter-rater reliability.^5^ Presumably as a result of inadequate syndrome awareness and barriers to diagnostic procedures (i.e. extended video-EEG), delay of diagnosis and treatment is common^6^ and contributes to avoidable adverse neurodevelopmental outcomes.^7,8^

Given these challenges, there is substantial need for novel diagnostic tools which may facilitate accurate and rapid diagnosis, or even allow detection of emerging IESS before clinical onset. Machine learning, especially deep learning, is a powerful data analysis technique that has been successfully used in the analysis of EEG. Deep learning algorithms have been developed to recognize and differentiate subtle morphological EEG features, and may even outperform human experts.^9^ In this study, we set out to determine whether a deep learning approach using only brief samples of interictal EEG could be used to accurately discriminate children with ES from normal controls.

## Methods

### Standard protocol approvals

This study was approved by the institutional review board at the University of California, Los Angeles. Given the retrospective design and deidentification of EEG data, the requirement for informed consent was waived.

### Study design and subjects

This is a retrospective cohort study, utilizing the same subjects and EEG samples that were used in our prior studies evaluating candidate biomarkers. Our methods for subject selection are described in detail therein.^10,11^ Briefly, our cohort consisted of 50 patients with epileptic spasms (cases) and 50 normal controls. All cases underwent overnight video-EEG which demonstrated epileptic spasms. The normal controls all underwent overnight video-EEG for evaluation of suspected seizures, but all normal controls were found to have events that were not seizures (e.g., gastoesophageal reflux, normal movements, etc.) and were deemed neurologically normal on a clinical basis.

### Data acquisition

Our methods for EEG data collection have been described previously.^10,11^ In brief, all clinical data were obtained from the electronic medical record. For each study, four EEG clips were abstracted (two in wakefulness and two in sleep, approximately 10 each). As such, a total of 400 EEG samples were used in the analyses described here. Importantly, the specific epochs selected for clipping were dictated by a randomization algorithm such that the abstractor selected EEG data beginning at a specific randomly selected time.

### EEG preprocessing and feature extraction

The overall methodologic framework is illustrated in Figure 1. EEG data from electrodes labeled Fp1, Fp2, F3, F4, C3, C4, P3, P4, O1, O2, F7, F8, T3, T4, T5, T6, FZ, CZ, and PZ were used. After applying a bandpass [1 Hz, 50 Hz] filter, EEG samples were segmented into windows of 15-seconds width. We applied the Fast Fourier Transform (FFT) to each channel to create a 2D channel-frequency image for each window. The input of the deep learning model is the 2D channel-frequency image of size 19 × 15000 (channels x frequency bins), with normalization to a standard gray scale image with pixel values ranging from 0 to 1.

**Figure 1.**
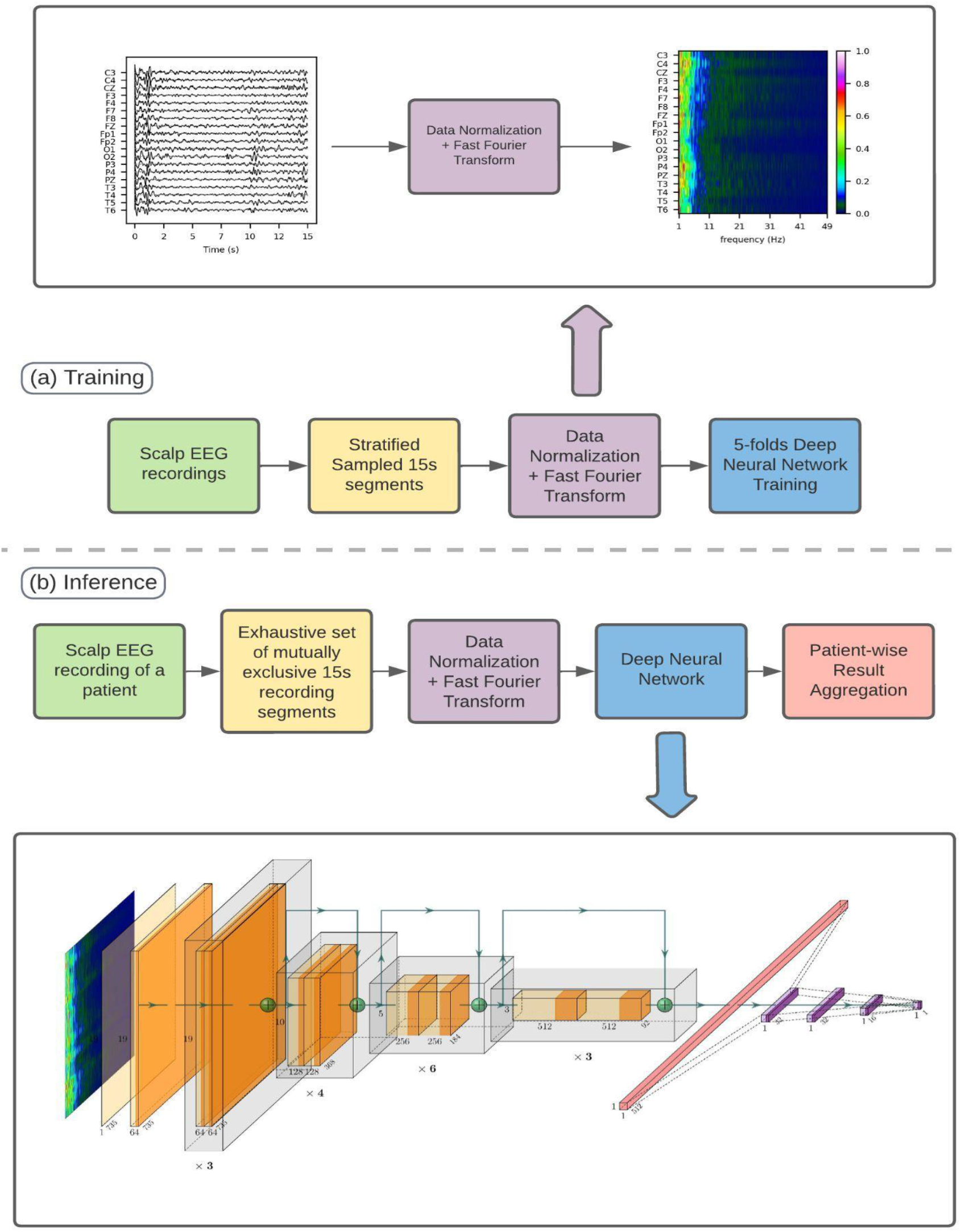
Methodologic Framework. The workflow consists of three parts. In part (i), each EEG recording is segmented into consecutive 15- second windows, which window then converted into a normalized FFT map. In part (ii), the FFT maps are used in 5-fold model training. In part (iii), the model inference procedure is illustrated, whereby non-overlapping 15-second segments are systematically sampled from each test recording and entered into the trained neural network. Subsequently, the results for each 15-second segment are consolidated to generate a classification outcome for the patient.

### Deep learning model architecture

To develop our model, we utilized the convolutional neural network (CNN), a state-of-the-art deep learning architecture that has demonstrated superior performance in the analysis of imaging^12^ and EEG data.^9^ To mitigate common gradient vanishing and facilitate superior model convergence, we used a 34- layer CNN with residual connections block. The output of the model is the probability of each EEG recording having been derived from a case.

### Model training and evaluation

Because ground truth labels (case/control) are missing for each 15s window of the EEG segment despite the availability of subject information, we adopted the weakly supervised learning approach, i.e. all windowed samples from case patients were labeled as case and control as control. We utilized a 5-fold cross-validation in which subjects were divided into five mutually exclusive groups, with 20 subjects (10 cases and 10 controls) in each group. For each fold, 80 subjects were used for model training and 20 subjects were used as a test set. Among the 80 training subjects, 10 patients (5 cases and 5 controls) were used for validation purposes to assess training performance in each training iteration. Overall, this training/evaluation approach supports the practical application of the model to novel subjects. Since the model is required to predict whether each 15-second EEG window is derived from a case or control, we used binary cross-entropy as the loss function, L=−[y·log(x)+(1−y)·log(1−x), where x is the probability of the windowed sample from the network, and the y is the label indicating case (positive, y = 1), or control (negative, y = 0). Given that EEG samples were of varying lengths, we adopted a novel stratified training approach which is widely used in the processing time-series data in the multimedia community.^13^ In each training iteration, for each patient, we randomly selected starting time stamps and constructed FFT maps from 15 second windows, such that information contributed from each patient was equal in each training iteration, with positive and negative samples balanced during the training phase. We conducted 400 training iterations in each fold. To improve generalization, we selected the model corresponding to the global minima in the validation set over training iterations, i.e., the highest balanced accuracy score.

As it is impractical to exhaustively evaluate model prediction on the FFT maps starting at every data point in the recording, we used a non-overlapping construction, such that FFT maps were generated from consecutive non-overlapping 15-second windows. To evaluate model performance we calculated accuracy, specificity, sensitivity, and F1 score. Finally, in order to classify a subject as case or control, we used the number of median-filtered successive case predictions in the non-overlapping construction, based on model confidence scores (predicted probability of each non-overlapping window). As such, a subject was predicted to be a case if any subset of windowed samples generated a prediction of case status. To protect external validity (i.e., generalization of the model to novel cohorts), we again employed a 5-fold cross-validation approach in the downstream analyses. Due to the sequential nature of the model inference (based on successive case predictions on the non-overlapping construction), we defined the earliest detection latency as the time from the beginning of each recording to the time when the model had sufficient information to classify the entire sample as having been derived from a case patient.

### Occlusion sensitivity analysis and salient band discovery

We next employed occlusion sensitivity analysis, a contemporary technique used in the interpretation of black-box classifiers such as CNN, in the attempt to identify the frequency band(s) in which critical EEG features may reside. Specifically, for each input column (frequency band) of an input image x, we evaluated the model with that frequency band missing and observed how the output (prediction) changed. In particular, a column (frequency band) is deemed important when the change of the model confidence ||f(x) - f(x_without_i)|| is large. The occlusion analyses were carried out on sleep and awake EEG models separately, using data samples from the non-overlapping construction. An occluded rectangular region (occlusion map) was defined as 19 channels x 5 Hz with a stride size of 1/15 Hz. We then generated a patient-wise occlusion template using the median value across each pixel in all patient-specific occlusion maps, and finally generated an overall sleep occlusion template across subjects by using the median value of each pixel across all patient-specific occlusion templates (n = 50). We repeated the same procedure for awake EEG to generate an overall awake occlusion template. The salient frequency band which contributes the most to the case prediction of the model was then identified using adaptive thresholding (Otsu’s method) in the overall occlusion template map.

### Targeted input perturbation

As illustrated in Figure 2, after salient frequency band identification by occlusion sensitivity analysis, the model was reassessed with targeted input perturbation, similar to methods previously employed by our group.^9,14^ As above, this procedure was conducted separately for awake and sleep data. For each 15 second window derived from Case EEG, a band-reject filter was applied to remove data within the salient frequency band, and these data were then replaced with average control data (average normalized waveform over the non-overlapping construction of all control patients) within the same frequency band. The effect of this perturbation across all case subjects was visually evaluated with histogram analysis of case/control predictions. Statistical assessment was accomplished with a one-tailed t-test performed on the change of output probability scores from the deep neural network.

**Figure 2.**
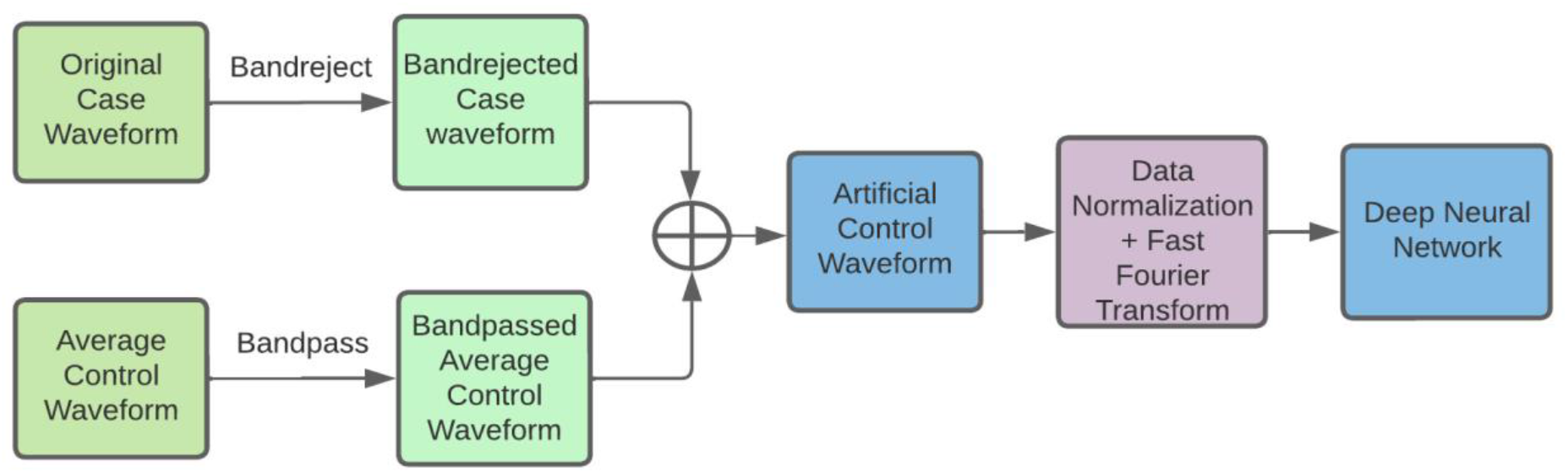
Targeted input perturbation workflow. The comprehensive workflow for implementing targeted input perturbation involves several steps. Initially, a specific 15-second segment of an EEG recording in waveform is chosen as the subject for perturbation. Frequencies other than the “biomarker” frequencies, which were determined from a previous occlusion sensitivity analysis, are retained by employing a band-reject filter. Following this, the so-called average control waveform is calculated by computing the numerical average waveform from all 15-second segment waveforms in control patients’ recordings. This average control waveform is then subjected to a band-pass filter to emulate normal control behavior within the biomarker band. The processed case waveform and average control waveform are subsequently combined to determine whether the case biomarker, as characterized by the processed case waveform, can transform the case waveform into a control. Finally, this synthesized waveform is input into the inference pipeline, and the resulting prediction from the deep neural network (DNN) is analyzed.

### Statistical Methods

All statistical calculations were accomplished with Python (v3.7.3; Python Software Foundation, USA). PyTorch (version 1.6.0; Facebook’s AI Research lab) was used to develop the deep neural network. Continuous variables are described by median (interquartile range) or mean (standard deviation). Comparisons between groups were performed using chi-square, t-test, or Wilxocon rank-sum as appropriate. Significant results were considered with p < 0.05 unless stated otherwise.

## RESULTS

### Model performance

The trained model performed satisfactorily with 5-fold cross validation. Using sleep and awake EEG data, respectively, accuracy in classifying cases and controls was 0.95 (SD 0.049) and 0.91 (0.041), recall was 0.96 (0.054) and 0.84 (0.11), precision (sensitivity) was 0.94 (0.052) and 0.98 (0.040), and F1 score was 0.95 (0.049) and 0.90 (0.052). In the final model, we enacted a threshold classifier such that a full-length EEG sample was predicted to have been derived from a case (patient) if any 6 consecutive 15 second windows were classified as “case”. With EEG preprocessing, the minimum sample required for case versus control prediction (i.e., detection latency) is 120 seconds. In patient-wise 5-fold cross-validation, the median detection latency was 120 seconds (95% CI 120 – 135).

### Interpretability analysis

In the occlusion sensitivity analysis, with the adaptive thresholding procedure, we found that model prediction was most affected by occlusion of data within the 9.7 – 22.0 Hz frequency band in sleep EEG, and the 1.0 – 8.6 Hz frequency band in awake EEG. After targeted input perturbation, in which these salient frequency bands were replaced with average control data, we found that patients who had been correctly classified as cases were now consistently re-classified as control subjects. The results of the targeted input perturbation are summarized in Figure 3.

**Figure 3.**
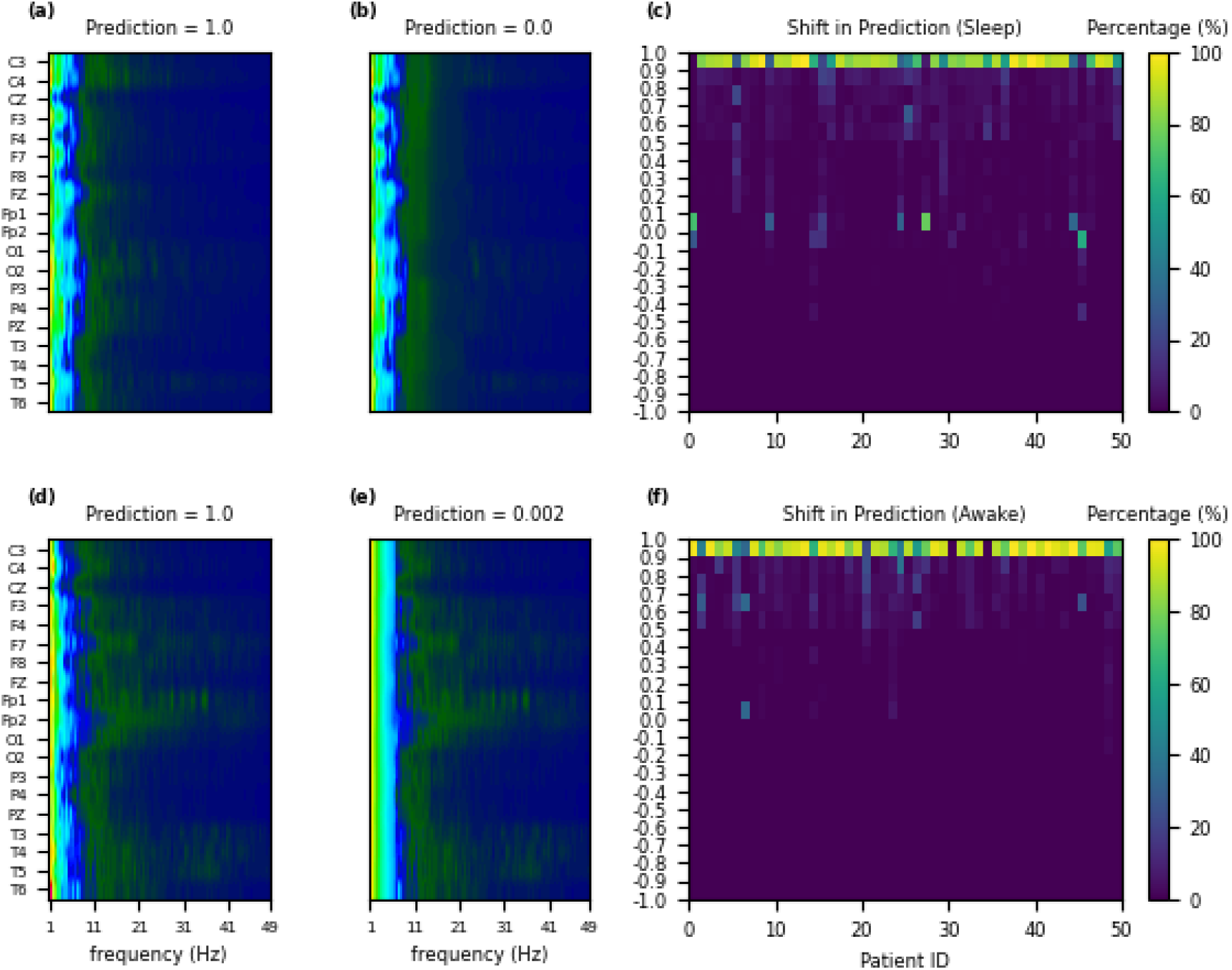
Targeted Input Perturbation. The top (a – c) and bottom (d – f) rows refer to sleep and awake datasets, respectively. Panels (a) and (d) illustrate sampled FFT maps from correctly classified cases. Panels (b) and (e) illustrate FFT maps after perturbation. Panels (c) and (f) illustrate the absolute shift in case prediction with perturbation.

## Discussion

In this study, we have demonstrated that a deep learning model can be successfully trained to accurately and efficiently distinguish children with epileptic spasms from normal controls, using relatively brief samples of interictal EEG. However, although our model is highly accurate, it is not altogether clear which attribute(s) the model is using to classify subjects. Our occlusion sensitivity and targeted input perturbation analyses suggest that the canonical beta frequency band is most critical for classification of sleep data, and that delta and low theta activity (1 to 6 Hz) is most important for evaluation of awake data. To a large extent, based on our visual review of extracted salient features, we suspect that the model is identifying cases on the basis of paroxysmal fast activity^15^ in sleep, and slowing or spike-slow-wave complexes in wakefulness.

There are several specific situations in which this model and approach may be useful. First, in clinical settings in which there is access to routine—but not extended video-EEG—this algorithm may enhance the diagnostic value of a brief routine EEG which does not capture ictal events. Second, to the extent that this model ‘detects’ IESS, this approach could be applied to augment EEG based surveillance for impending epileptic spasms relapse after initially successful treatment, or to screen populations at risk of developing IESS (e.g., Tuberous Sclerosis Complex or hypoxic ischemic encephalopathy).

There are several significant limitations to this study. Foremost, this is a retrospective analysis and a prospective external validation study is needed. In addition, although the sample size (n = 100) is relatively large for a study of children with epileptic spasms, it is rather small from the perspective of machine learning and deep learning. A much larger sample may slightly improve classification accuracy—given that there is limited room for improvement—and may significantly improve identification of salient frequency bands and facilitate discovery of novel EEG features. A disappointing aspect of this initial analysis was the lack of discovery of a novel EEG biomarker.

In summary, this is a successful first attempt to train an EEG- based deep learning model to distinguish children with epileptic spasms from normal controls. Further study is needed to validate our findings, and confront more challenging clinical questions such distinguishing children with epileptic spasms from children with other seizure types, or using EEG to predict emergence of IESS, response to treatment, relapse, and potential evolution to Lennox-Gastaut Syndrome.

## Data Availability

All data produced in the present study are available upon reasonable request to the authors

## Acknowledgements

This study was accomplished with generous support from the John C. Hench Foundation.

## Disclosure of Conflicts of Interest

Dr. Daida has received research support from the Uehara Memorial Foundation and the SENSHIN Medical Research Foundation.

Dr. Rajaraman has received research support from Marinus Pharmaceuticals, GW Pharmaceuticals, the Pediatric Victory Foundation, and the International Foundation for CDKL5 Research (IFCR).

Dr. Nariai is supported by the National Institute of Neurological Disorders and Stroke (NINDS) K23NS128318, the Sudha Neelakantan & Venky Harinarayan Charitable Fund, the Elsie and Isaac Fogelman Endowment, and the UCLA Children’s Discovery and Innovation Institute (CDI) Junior Faculty Career Devel-opment Grant (#CDI-TTCF-07012021).

Dr. Hussain has received research support from the John C. Hench Foundation, the CJDA Foundation, the Mohammed F. Alibrahim Endowment, the Elsie and Isaac Fogelman Endowment, the Epilepsy Therapy Project, the Milken Family Foundation, Paul Hughes Family Foundation, the Pediatric Epilepsy Research Foundation, Eisai, Bio-Pharm, Lundbeck, Insys, GW Pharmaceuticals, UCB Biopharma, Zogenix, Marinus, and the NIH. He has received compensation for service as a consultant to Amzell, Aquestive Therapeutics, Equilibre Biopharmaceuticals, Insys, GW Pharmaceuticals, Mallinckrodt, Marinus, MGC Pharmaceuticals, Radius, Shennox, UCB Biopharma, Upsher-Smith Laboratories, West Therapeutic Development, and Zogenix.

The remaining authors report no conflicts of interest.

## Notes

### Funding Statement

This study was supported by the John C. Hench Foundation.

### Author Declarations

The IRB of the University of California, Los Angeles, gave ethical approval for this work.

## References

1. Gaily E, Lommi M, Lapatto R, Lehesjoki A-E. Incidence and outcome of epilepsy syndromes with onset in the first year of life: A retrospective population-based study. Epilepsia. 2016;57:1594–1601.

2. Grinspan ZM, Mytinger JR, Baumer FM, Ciliberto MA, Cohen BH, Dlugos DJ, Harini C, Hussain SA, Joshi SM, Keator CG, Knupp KG, McGoldrick PE, Nickels KC, Park JT, Pasupuleti A, Patel AD, Shahid AM, Shellhaas RA, Shrey DW, Singh RK, Wolf SM, Yozawitz EG, Yuskaitis CJ, Waugh JL, Pearl PL. Management of Infantile Spasms During the COVID-19 Pandemic. J Child Neurol. 2020;35:828–834.

3. Specchio N, Pietrafusa N, Ferretti A, De Palma L, Santarone ME, Pepi C, Trivisano M, Vigevano F, Curatolo P. Treatment of infantile spasms: why do we know so little? Expert Rev Neurother. 2020;20:551–566.

4. Caraballo RH, Fortini S, Reyes G, Carpio Ruiz A, Sanchez Fuentes SV, Ramos B. Epileptic spasms in clusters and associated syndromes other than West syndrome: A study of 48 patients. Epilepsy Research. 2016;123:29–35.

5. Hussain SA, Kwong G, Millichap JJ, Mytinger JR, Ryan N, Matsumoto JH, Wu JY, Lerner JT, Sankar R. Hypsarrhythmia assessment exhibits poor interrater reliability: A threat to clinical trial validity. Epilepsia. 2015;56:77–81.

6. Hussain SA, Lay J, Cheng E, Weng J, Sankar R, Baca CB. Recognition of infantile spasms is often delayed: The ASSIST study. J Pediatr. 2017;190:215–221.

7. O’Callaghan FJK, Lux AL, Darke K, Edwards SW, Hancock E, Johnson AL, Kennedy CR, Newton RW, Verity CM, Osborne JP. The effect of lead time to treatment and of age of onset on developmental outcome at 4 years in infantile spasms: evidence from the United Kingdom Infantile Spasms Study. Epilepsia. 2011;52:1359–1364.

8. Auvin S, Hartman AL, Desnous B, Moreau A-C, Alberti C, Delanoe C, Romano A, Terrone G, Kossoff EH, Del Giudice E, Titomanlio L. Diagnosis delay in West syndrome: misdiagnosis and consequences. Eur J Pediatr. 2012;171:1695–1701.

9. Zhang Y, Lu Q, Monsoor T, Hussain SA, Qiao JX, Salamon N, Fallah A, Sim MS, Asano E, Sankar R, Staba RJ, Engel J, Speier W, Roychowdhury V, Nariai H. Refining epileptogenic high-frequency oscillations using deep learning: a reverse engineering approach. Brain Commun. 2022;4:fcab267.

10. Smith RJ, Hu DK, Shrey DW, Rajaraman R, Hussain SA, Lopour BA. Computational characteristics of interictal EEG as objective markers of epileptic spasms. Epilepsy Res. 2021;176:106704.

11. Miyakoshi M, Nariai H, Rajaraman RR, Bernardo D, Shrey DW, Lopour BA, Sim MS, Staba RJ, Hussain SA. Automated preprocessing and phase-amplitude coupling analysis of scalp EEG discriminates infantile spasms from controls during wakefulness. Epilepsy Research. 2021;178:106809.

12. He K, Zhang X, Ren S, Sun J. Deep Residual Learning for Image Recognition. 2016 IEEE Conference on Computer Vision and Pattern Recognition (CVPR). 2016. p. 770–778.

13. Lu Q, Zhang Y, Lu M, Roychowdhury V. Action-conditioned On-demand Motion Generation. Proceedings of the 30th ACM International Conference on Multimedia [Internet]. New York, NY, USA: Association for Computing Machinery; 2022 [cited 2023]. p. 2249–2257. Available from: https://dl.acm.org/doi/10.1145/3503161.3548287

14. Zhang Y, Chung H, Ngo JP, Monsoor T, Hussain SA, Matsumoto JH, Walshaw PD, Fallah A, Sim MS, Asano E, Sankar R, Staba RJ, Engel J, Speier W, Roychowdhury V, Nariai H. Characterizing physiological high-frequency oscillations using deep learning. J Neural Eng. 2022;19:1–17.

15. Wu J, Koh S, Sankar R, Mathern GW. Paroxysmal fast activity: an interictal scalp EEG marker of epileptogenesis in children. Epilepsy Res. 2008;82:99–106.

